# Prevalence and determinants of evidence of silicosis and impaired lung function, among small scale tanzanite miners and the peri-mining community in northern Tanzania

**DOI:** 10.1101/2023.12.13.23299915

**Authors:** Alexander W. Mbuya, Innocent B. Mboya, Hadija H. Semvua, Simon H. Mamuya, Patrick J. Howlett, Sia E. Msuya

**Affiliations:** Department of Community Health, Institute of Public Health, Kilimanjaro Christian Medical University College, Moshi, Tanzania; Department of Community Health, Kibong’oto Infectious Diseases Hospital, Kilimanjaro, Tanzania; Department of Epidemiology and Biostatistics, Kilimanjaro Christian Medical University College, Moshi, Tanzania; Kilimanjaro Clinical Research Institute, Kilimanjaro Christian Medical Center, Moshi, Tanzania; Department of Environmental and Occupational Health, School of Public Health and Social Sciences, Muhimbili University of Health and Allied Sciences, Dar es Salaam, Tanzania; National Heart & Lung Institute, Imperial College London, Guy Scadding Building, Cale Street, London, SW3 6LY; Department of Community Health, Kilimanjaro Christian Medical Center, Moshi, Tanzania

**Keywords:** silicosis, impaired lung function, chronic obstructive pulmonary diseases, restrictive lung diseases, miners, small-scale miners, tanzanite, Tanzania

## Abstract

Among mining communities in Tanzania, the limited data available suggests the prevalence of silicosis, obstructive lung diseases (OLD) and restrictive lung disease (RLD) to be around 1.6%, 1.9% and 8.8% respectively. Our study therefore aimed to determine the prevalence and factors associated with evidence of silicosis and ILF among tanzanite mining community in northern Tanzania.

We conducted a cross-sectional study, involving 330 randomly selected miners and 330 conveniently selected non-mine workers from the peri-mining community (PMC) in Mererani mines, northern Tanzania. Evidence of silicosis was defined based on study participants’ history of exposure to mining dust and digital chest radiological findings with reference to the 2011 ILO classification of pneumoconiosis. Impaired lung function was determined by spirometry using American Thoracic Society (ATS)/European Respiratory Society (ERS) recommended system 3. Association between evidence of silicosis/impaired lung function and presumed risk factors were determined using binary logistic regression analyses. The study found that 99/330 (30.0%) of miners had evidence of silicosis, of whom 97.0% had accelerated silicosis. Among miners and community members, 75 (11.4%) had ILF, of whom 30 (4.5%) had COPD, 9 (1.4%) had asthma, 29 (4.4%) had restrictive lung disease and 7 (1.1%) had mixed pattern of both obstructive and restrictive lung disease. We found that having a daily income of more than USD 4.3 was associated with lower odds of silicosis (aOR 0.57, 95% CI 0.37-0.89, p<0.05) while ILF was associated with being a miner (aOR 2.06, CI=1.38-3.07, p<0.001).

We found a concerningly high prevalence of evidence of silicosis despite short durations of exposure among small scale tanzanite miners. Immediate dust control measures including deployment of wet drilling, wearing of personal protective equipment (PPE) and regular monitoring of dust exposure need to be enforced by the OSHA (Tanzania).

## Introduction

Silicosis is an ancient, progressive, incurable, but potentially preventable chronic lung disease caused by the inhalation of crystalline silica. The disease occurs following exposure to respirable crystalline silica (RCS) mostly in the mining and manufacturing industries. Three forms of silicosis are described although not distinctly defined. Acute silicosis, the most histopathologically distinct, may occur following between a few months and up to 5 years of heavy exposure to RCS. Accelerated silicosis occurs over 1-10 years with chronic silicosis occurring over longer periods [1]<u>–</u>[3]. In majority of mining populations, acute silicosis and accelerated disease may be considered rare. Study findings among South Africa (SA) gold miners showed the crude prevalence of chronic silicosis to range between 1.8% to 8.6% [4]. Data on silicosis in Tanzania are scarce but the prevalence among miners is estimated at 1.6% [5]. Majority of studies, however, are based in large scale miners (LSM) populations. The SSM engage in extraction from ore or mineral deposits using low-impact, potentially short-term, small-footprint, regulated mining operations and technologies that are usually not labour-intensive [6], though sometimes they may be similar to Artisanal Small Scale Miners (ASM) by being labour intensive. Studies of SSM have suggested that prevalence may be up to 29.1% with shorter exposure periods [7], [8]. A recent systematic review found a prevalence of between 11 and 37% among small-scale miners [9]. Previous evidence of very high silica exposure has been documented in Mererani [10], [11] reflecting simple mechanization comprising of dry pneumatic drills, air blowers, and explosives, with almost no form of dust reduction measures such as wet drilling being deployed or use of PPE. Despite this, the prevalence of silicosis amongst tanzanite miners is unknown.

Tuberculosis is common among mining communities, and silicosis has been shown to significantly increase the risk of tuberculosis, particularly in the context of HIV [12], [13]. Little is known about tuberculosis rates among SSM, however some evidence available from Ghana suggests the prevalence is high [14] and also recent report among SSM in Mererani has shown the prevalence to be around 7.0% [15]. Both OLD and RLD are associated with occupational dust exposure [16]. The prevalence of Chronic Obstructive Pulmonary Diseases (COPD) among individuals exposed to dust in China has been reported to be 32.7% and was noted to be highest among those diagnosed to have silicosis [17]. Underground gold miners in Tanzania have been reported to have OLD prevalence of 1.9% and RLD prevalence of 8.8% [18].

Based on the evidence of the occupational risks among the SSM and the clinical experience of the authors, this study aimed to determine the prevalence and factors associated with evidence of silicosis and ILF among tanzanite mining community in northern Tanzania. The aim of the study has been well achieved.

## Materials and methods

### Study design and study site

We have previously described in details our study methods [15]. In brief however, we conducted a cross-sectional study in Mererani mines located in Simanjiro district, in northern Tanzania. Simanjiro District is estimated to have a population of 221,211 based on national census of 2012 and a population growth rate of 2.4% [19], with a little more than half being located in Mererani ward where the tanzanite mines are found. Mererani is the only place in the world where tanzanite gemstone is being mined at a commercial scale. Mererani town is located about 5 kilometres from the mines and 70 kilometres from Arusha City. The tanzanite mines in Mererani are all fenced with only one, guarded entrance gate. The weather in Mererani area is generally dry and strong winds with frequent change of directions. For this reason, as well as a control group, we chose to evaluate the PMC for dust related diseases.

### Study population

The study recruited men aged 18 years and above from two groups: SSM and PMC members. Miners were defined as an individual who goes down the mining pits and engages in various activities including drilling, blasting and shovelling of rocks. As for this study, only SSM engaging with drilling activities were involved, though there is frequent task shifting in which any miner can perform any of the different activities. The PMC were residents of Mererani town engaging in other non-mining activities who have no history of working in the mines. Most miners work shifts lasting between 10 to 12 hours, but some may go up to 48 hours. Our experience is that all underground miners are men; hence only men were recruited as study participants. Miners in Mererani utilize simple mechanization using handheld pneumatic drills, wheelbarrows and sacks, and diesel-based pulley systems. At the time of data collection, there were 100 active mining pits, each with between 70 to 90 SSM, hence a maximum of 9,000 active SSM.

### Sample size

Based on 16% estimated (from routine clinical practice) prevalence of silicosis among tanzanite miners in Tanzania, the minimum sample size of 206 participants were selected from the SSM group based on the below formular:

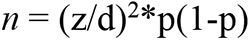

whereby:

*n* = minimum sample size

z = critical value for 95% confidence interval (1.96)

d = degree of precision (0.05)

p = estimated prevalence of silicosis (16%)

This led to *n* = 206.4, which was then adjusted by +10% for lung function assessment, +10% for TB investigation and addition +30% for data loss, hence total sample came to 325, which was approximated to 330 SSM. The same number was taken for the PMC to make the overall total sample of 660.

### Sampling and data collection procedures

Following ethical clearance we introduced the study to mine managers, who agreed to participate. We randomly sampled 22 pits from a coded list of the 100 mining pits. From each selected pit, a coded list of all mine workers was obtained forming the sampling frame. To attain a sample size of 330, and to avoid disruption of ongoing work, we used random sampling to select 15 SSM from each pit. This number of 15 SSM per pit ensured a reasonable number of workers remained to continue with work while the selected ones visited the clinic for disease evaluation.

For the PMC, a list of 85 registered streets in Mererani town obtained from the local government authority. From this, a random sample of 22 streets was selected and, in each street, a reference house was established at the street’s main junction with the Kilimanjaro International Airport (KIA) to Mererani highway. Houses (being it a shop, health facility or school) were selected consecutively, and from each selected house, individual participants were selected conveniently (as per inclusion and exclusion criteria) until 15 participants per street (cluster) were reached, making a total of 330 participants. For purposes of comparison with the miners, only men were selected as study participants.

Participants were scheduled into groups of 20 up to 25 individuals and provided with return transport to the Occupational Health Service Center (OHSC) at Kibong’oto Infectious Diseases Hospital (KIDH), located about 60 kilometres from Mererani town. Procedures performed to the study participants at the OHSC included taking each participant through the questionnaire adapted from Medical Research Council (MRC) United Kingdom (UK) Respiratory Questionnaire (MRC, 1986), digital chest X-ray examination (DRGEM Corporation, Korea) and spirometry test using Easy on-PC (NDD Medizintechnik AG, Zurich, Switzerland). The study radiologist (based at KIDH) with extensive experience of interpreting digital chest radiograph of pneumoconiosis read and interpreted the chest X-rays of the study participants based on the 2011 ILO Pneumoconiosis Classification System with a 12-points scale (4 categories and 12 sub-categories). In the absence of the ability to histopathologically confirm acute silicosis and in keeping with previous research [8], in those with chest radiological evidence of silicosis, participants with less than 15 years of exposure were classified as having accelerated silicosis and those with at least 15 years of exposure as having chronic silicosis.

The spirometer was calibrated twice per day, using a 3L Volume Calibration Syringe (CRC Medical, DMS Limited, UK). Participants with initial obstructive spirometry results were tested for bronchodilator reversibility. An increase of at least 200mls in FEV_1_ following the bronchodilator reversibility test was regarded as a significant response, hence was diagnosed as asthma. Participants who failed to perform the initial spirometry test or could not produce reliable results, were re-scheduled for other examinations in one-week intervals until reliable results were obtained, meaning achieving the acceptability and repeatability criteria. Acceptability was done by evaluating each blow curve i.e. flow-volume and volume-time graphs for good quality. This include a sharp start with no hesitation, maximum inspiration and expiration, absence of air flow cessation, absence of cough, inspiration during the trace & leaks and exhalation lasting for at least 6 seconds with less than 50mls being exhaled in the last 2 seconds. Repeatability was assessed by checking (from tests that have achieved the acceptability criteria) the best two values of FEV_1_ and FVC to be within 150mls of each other and if FVC is <1litre, then the best values of FEV_1_ and FVC to be within 100mls of each other [20].

Importantly, unlike in routine clinical practice in which only presumptive (those with symptoms) TB cases are asked to produce sputum, in our study all participants were asked to produce sputum for TB investigation, with a positive GeneXpert MTB/RIF (GeneXpert Dx System, Version 4.8, Cepheid, USA) defining a diagnosis for TB. HIV rapid blood tests (SD BIOLINE HIV-1/2 3.0 Standard Diagnostic Inc.) and fasting blood glucose tests (GlucoPlus^TM^, GlucoPlus Inc. Quebec, Canada) were performed. Participants found to have any of the investigated diseases, were enrolled to care and treatment services as per the Ministry of Health (MoH) guidelines.

### Data management and analysis

Paper case report forms were entered into Excel. Analysis was performed using Statistical Package for Social Sciences (SPSS) software (IBM SPSS Statistics Version 27). The primary outcomes were presence/absence of evidence of silicosis and presence/absence of ILF while the secondary outcomes were the forms of silicosis (whether it is accelerated or chronic based on the duration of occupational exposure to RCS among those with evidence of silicosis) and the grades of COPD. As per this paper, silicosis was diagnosed based on two key factors i.e. history of a participant being exposed to occupational silica dust and having radiological digital chest X-ray findings consistent with silicosis as diagnosed by a radiologist experienced in this field. The PMC was also assumed to be exposed to the mining dust based on being near to the mines, the practice of leaving the mined rocks above ground in open air and given the environmental characteristics of few vegetation and frequent winds. Impaired lung function comprised of three disease conditions: Obstructive lung disease, restrictive lung disease and mixed (obstructive and restrictive) lung disease. Asthma was diagnosed as an increase of at least 120mls in either of FEV_1_ or FVC following bronchodilator reversibility tests among those found to have obstructive lung disease while those with no response/or an increase of less than 120mls in either FEV_1_ or FVC on bronchodilator reversibility test, were classified as having COPD.

The ATS/ERS recommended system 3 based on defining airways obstruction using FEV_1_/FVC < Lower Limit of Normal (LLN) and using Z scores for FEV_1_ to classify severity (grades 1 to 5) for COPD were used in this study. The LLN Z-score of -1.64 was used for diagnosis of both COPD, RLD and Mixed COPD/RLD patterns, while for COPD and restrictive diseases grading the FEV_1_ Z-scores used were: Z-score ≥ -2 (grade 1), -2.5 ≤ Z-score < -2 (grade 2), -3 ≤ Z-score < -2.5 (grade 3), -4 ≤ Z-score < -3 (grade 4) and Z-score < -4 (grade 5).

The association between evidence of silicosis, our primary outcome, and presumed risk factors was determined using a priori logistic regression analysis. The key explanatory variables for inclusion in both models were duration of work in Mererani, history of previous lung disease, TB, smoking and income level. A similar model was developed for impaired lung function. A statistically significant level of p<0.05 was used while p<0.001 was considered as statistically highly significant.

### Ethical consideration

Ethical clearance was obtained from the Kilimanjaro Christian Medical University College (KCMUCo) No. 2416 and the Medical Research Coordinating Committee of the National Institute for Medical Research (NIMR) No: NIMR/HQ/R.8a/Vol.IX/3308. Permission to conduct the study was sought from the Permanent Secretary – Presidents’ Office, Regional Administration & Local Government (PS-PORALG), and the owners/managers for the specific mining pits. Written informed consent, translated in Kiswahili language was obtained from each study participant.

## Results

### Demographic and clinical characteristics

Table 1 shows the socio-demographic and clinical characteristics of the mining communities in Mererani. The SSM comprises of relatively younger population with a median age of 35 years, compared to the PMC median of 39 years. The SSM have significantly higher proportion of individuals with lower education level compared to the PMC. The SSM have significantly lower duration of working in Mererani with a median duration of 5 years compared to 8.5 years for the PMC. The SSM have a significantly lower median daily income of about USD 2.9 compared to the PMC median of USD 7.2. A higher proportion of the PMC (7.9%) were found to have TB compared 6.1% for the SSM while all (100%) of those with evidence of silicosis were SSM.

**Table 1.** Socio-demographic and clinical characteristics of the Mining Communities in Mererani (N=660)

| Characteristics | Total | Small Scale Miners<br>(n = 330) | Peri-Mining Community<br>(n = 330) |
| --- | --- | --- | --- |
| **Age (years)** |  |  |  |
| Median (IQR) | 38 (31.0-45.0) | 35.0 (30.0-44.0) | 39.0 (32.0-46.0) |
| 18-30 | 158 (23.9) | 92 (27.8) | 66 (20.0) |
| 31-40 | 254 (38.5) | 129 (39.1) | 125 (37.9) |
| 41-50 | 180 (27.3) | 84 (25.5) | 96 (29.1) |
| >50 | 68 (10.3) | 25 (7.6) | 43 (13.0) |
| **Marital Status** |  |  |  |
| Not in relationship | 112 (17.0) | 63 (19.1) | 49 (14.8) |
| In a relationship | 548 (83.0) | 267 (80.9) | 281 (85.2) |
| **Education level** |  |  |  |
| ≤Primary | 468 (70.9) | 272 (82.4) | 196 (59.4) |
| >Primary | 192 (29.1) | 58 (17.6) | 134 (40.6) |
| **Duration of work in Mererani (years)** |  |  |  |
| Median (IQR) | 7 (5-11) | 5.0 (4.0 – 7.0) | 8.5 (6.0 – 12.0) |
| ≤5<sup>a</sup> | 238 (36.0) | 179 (54.2) | 59 (17.9) |
| 6-10 | 254 (38.5) | 117 (35.5) | 137 (41.5) |
| >10 | 168 (25.5) | 34 (10.3) | 134 (40.6) |
| **Income (USD/day)** |  |  |  |
| Median (IQR) | 4.3 (2.8-8.6) | 2.9 (2.2-7.2) | 7.2 (4.3-11.5) |
| >4.3<sup>b</sup> | 432 (65.5) | 157 (47.6) | 275 (83.3) |
| ≤4.3 | 228 (34.5) | 173 (52.4) | 55 (16.7) |
| **Smoking status** |  |  |  |
| **Median duration** |  | 10.0 (4.0 – 15.8) | 11.0 (5.0 – 17.0) |
| **(years)** |  |  |  |
| Smokers | 93 (14.1) | 60 (18.2) | 33 (10.0) |
| Non-smokers | 567 (85.9) | 270 (81.8) | 297 (90.0) |
| **Previous Lung Diseases** |  |  |  |
| Yes | 351 (53.2) | 205 (62.1) | 146 (44.2) |
| No | 309 (46.8) | 125 (37.9) | 184 (55.8) |
| **Tuberculosis** |  |  |  |
| Yes | 46 (7.0) | 20 (6.1) | 26 (7.9) |
| No | 614 (93.0) | 310 (93.9) | 304 (92.1) |
| **Lung Function: median** |  |  |  |
| **(IQR)** |  |  |  |
| FEV<sub>1</sub> | 2.80 (2.27-3.36) | 2.85 (2.17-3.42) | 2.75 (2.29-3.31) |
| FVC | 3.61 (3.03-4.10) | 3.66 (2.97-4.15) | 3.53 (3.09-4.02) |
| FEV<sub>1</sub>/FVC | 0.80 (0.72-0.85) | 0.80 (0.71-0.85) | 0.81 (0.73-0.85) |
| **HIV status** |  |  |  |
| Positive | 8 (1.2) | 6 (1.8) | 2 (0.6) |
| Negative | 652 (98.8) | 324 (98.2) | 328 (99.4) |
| **Evidence of silicosis<sup>c</sup>** |  |  |  |
| Yes | 99 (15.0) | 99 (30.0) | 0 (0.0) |
| No | 561 (85.0) | 231 (70.0) | 330 (100.0) |

<sup>a</sup>Cut-off duration of exposure to RCS for patients with acute form of silicosis. As acute silicosis requires histopathological confirmation (in additional to the radiological chest findings) which was not done in this study, this group has been analysed under 'accelerated silicosis' i.e. exposure to RCS for <15years, <sup>b</sup>The overall median income of the study participants was USD 4.3 per day. <sup>c</sup>Evidence of silicosis refers to silicosis diagnosed based only on history of occupational exposure to RCS and chest radiological findings by digital X-ray.

### Evidence of silicosis and lung function status

Figure 1 shows the duration of work in Mererani mines among the SSM found to have evidence of silicosis in which round 97% falls under the working duration of less than 15 years, consistent with findings of accelerated silicosis.

**Figure 1.**
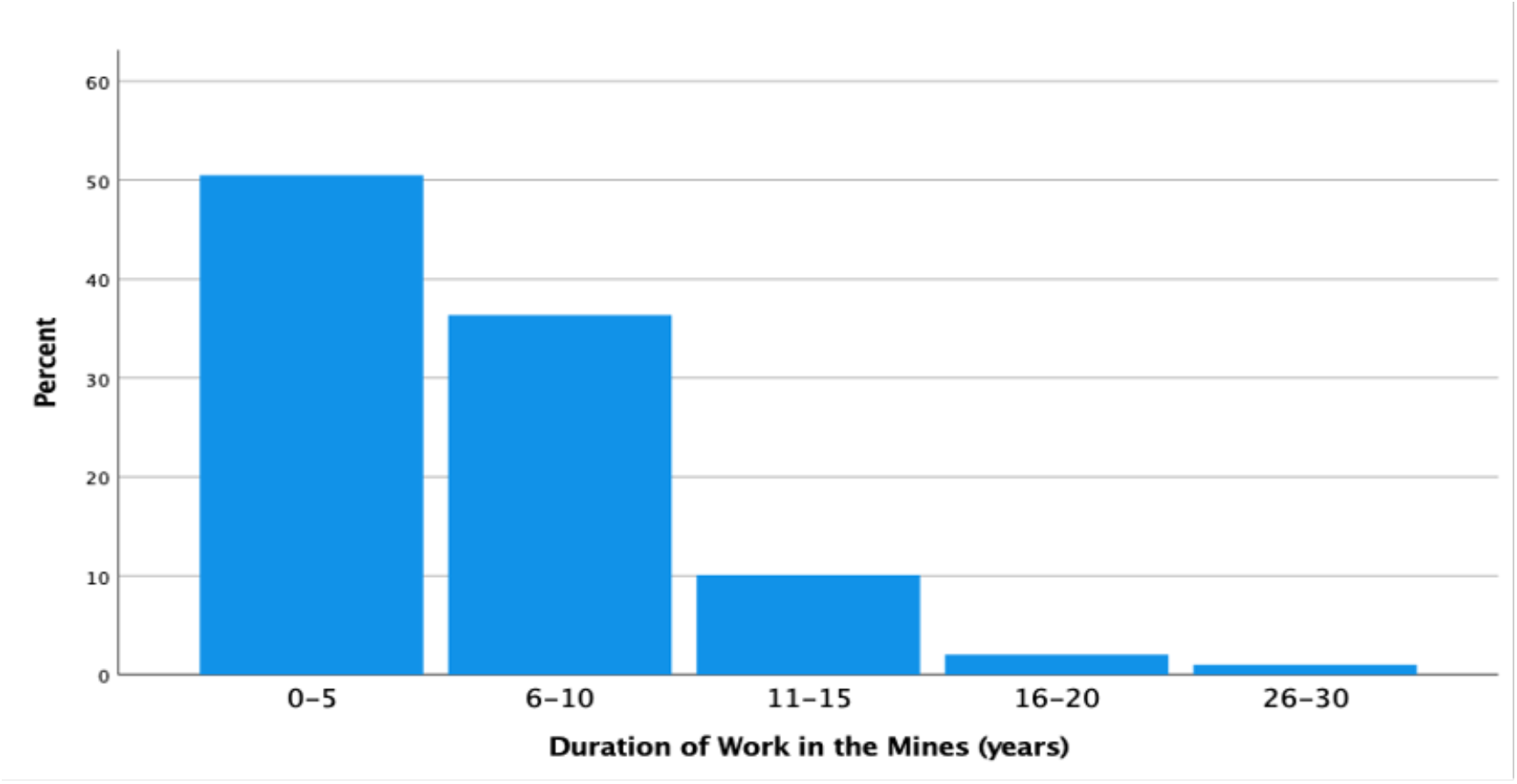
Duration of work in Mererani mines among mine workers found to have evidence of silicosis (N=99). Most miners (>50%) had a working duration of up to 5years while 97% had a working duration of less than 15years.

Among the 660 study participants, 75 (11.4%) were found to have various forms of ILF, in which 30 (40.0%) had chronic obstructive lung disease, 9 (12.0%) had asthma, 29 (38.7%) had restrictive lung disease and 7 (9.3%) had mixed pattern of both obstructive and restrictive lung disease. Of the 75 participants found to have ILF, 48 (64%) were SSM. Table 2 shows the lung function status among SSM and the PMC based on the LLN Z-score of -1.64 for FVC, FEV_1_ and FEV_1_/FVC and asthma for those with an increase of at least 120mls in either of FEV_1_ or FVC following bronchodilator reversibility tests among those found to have obstructive lung disease. Majority, 39 (52%) of the study participants with ILF were found to have obstructive lung disease, of whom 24 (61.5%) were SSM and 15 (38.5%) were from the PMC.

**Table 2.** Lung Function Status Among Mining Communities in Mererani (660)

| Lung Function Status | Total | Miners | Peri-Mining Community |
| --- | --- | --- | --- |
| Normal | 585 (100.0) | 282 (48.2) | 303 (51.8) |
| Restrictive | 29 (100.0) | 19 (65.5) | 10 (34.5) |
| Mixed | 7 (100.0) | 5 (71.4) | 2 (28.6) |
| Chronic obstructive | 30 (100.0) | 17 (56.7) | 13 (43.3) |
| Asthma | 9 (100.0) | 7 (77.8) | 2 (22.2) |

Figure 2 shows the scatter plot for the various forms of lung function status as function of FVC and FEV_1_. The pattern is almost linear with majority of FVC and FEV_1_ intercepts representing normal lung function findings (blue circles) belonging to the upper right half of the plot. Most of the rest of the circles for various forms of ILF are found on the lower left region of the scatter plot.

**Figure 2.**
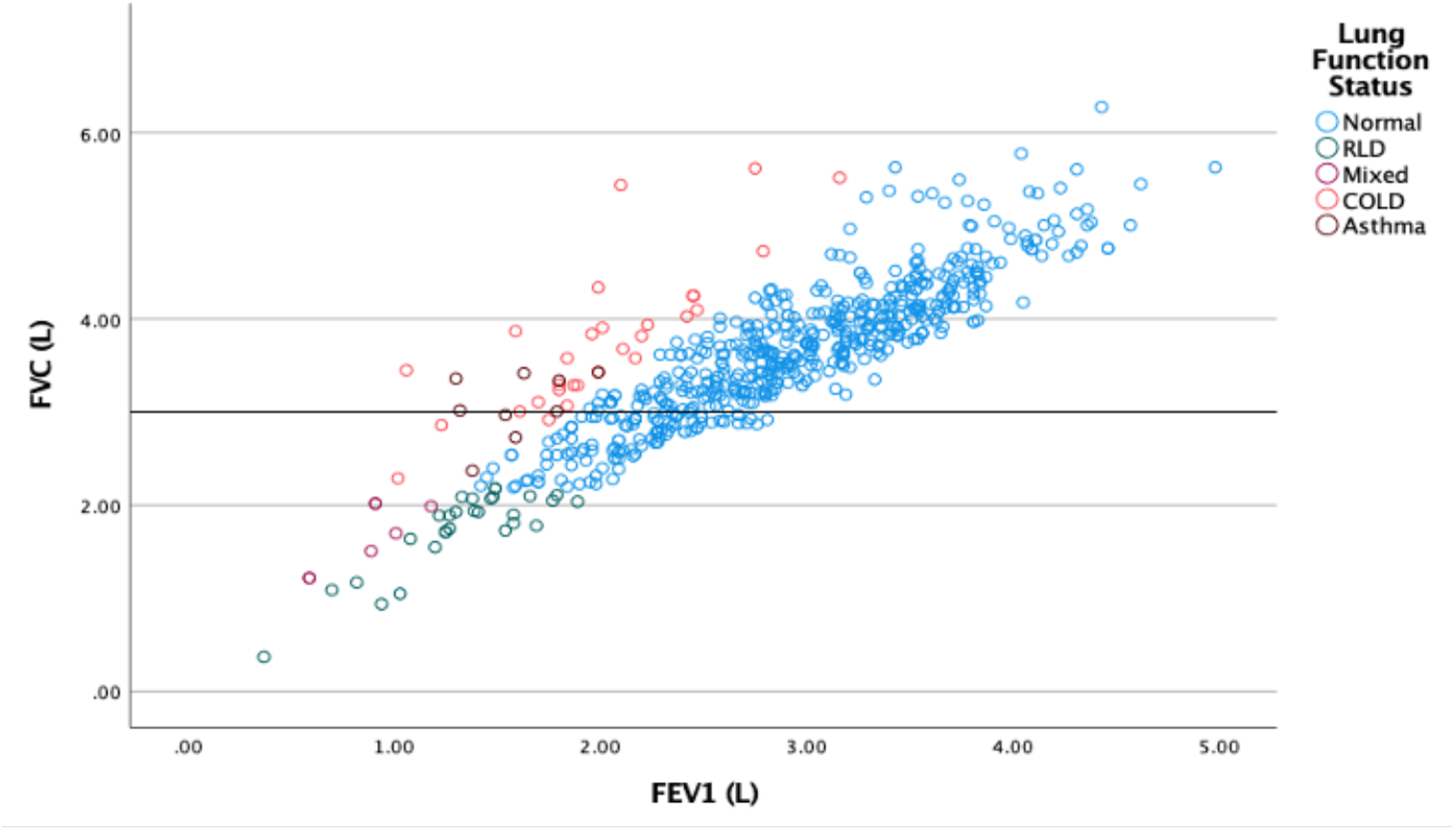
Scatter plot showing the spread of various forms of lung function status as functions of FVC and FEV_1_ (N=660)

Table 3 shows the grades (severity) of impaired lung function based on ATS/ ERS recommended system 3 using Z scores for FEV_1_. Majority 50 (84.7%) of those with ILF had mild (grade 1) disease, of whom 27 (54.0%) were miners and 23 (46.0%) were from PMC.

**Table 3.**
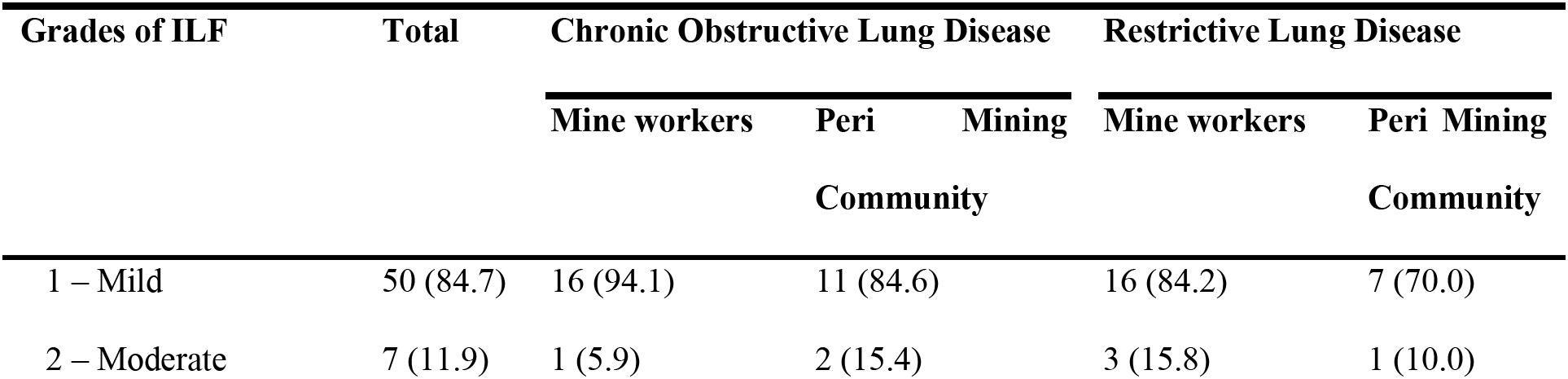
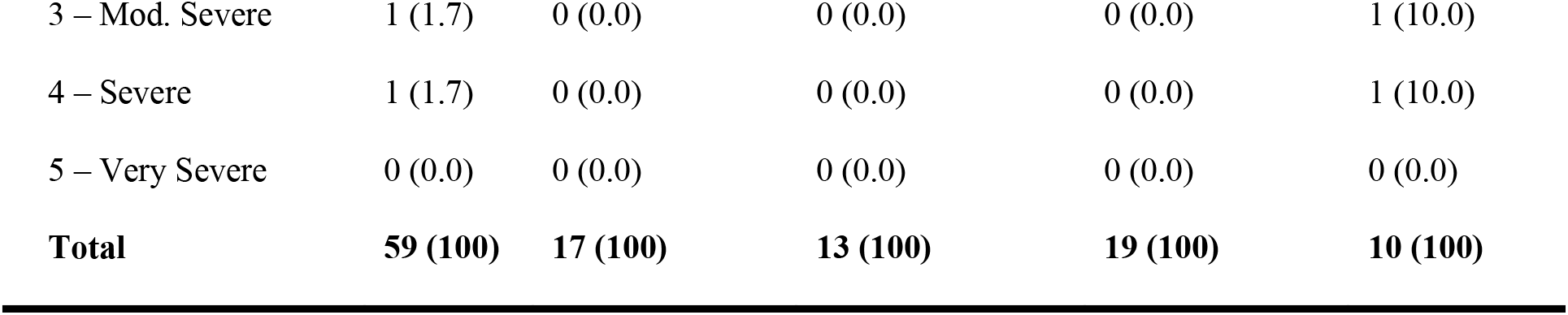
Grades of impaired lung function among those with COPD and RLD among mining communities in Mererani (N=59)

### Factors associated with evidence of silicosis

Crude and adjusted associations between a set of predictors and evidence of silicosis among the mining communities in Mererani are shown in table 4. Those earning above USD 4.3 per month had 43% lower odds of having evidence of silicosis (aOR 0.57, CI=0.37-0.89, p=0.013).

**Table 4.** Factors Associated with Evidence of Silicosis among Mining Communities in Mererani (N=660)*

| Variable | cOR (95%CI) | p-value | aOR (95%CI) | p value |
| --- | --- | --- | --- | --- |
| **Working duration in Mererani (years)** |  |  |  |  |
| <15 | 2.38 (0.93-6.07) | 0.070 | 2.04 (0.78-5.34) | 0.147 |
| ≥15 | Ref. |  | Ref. |  |
| **Income (USD/day)** |  |  |  |  |
| >4.3 | 0.50 (0.33-0.77) | 0.002 | 0.57 (0.37-0.89) | 0.013 |
| ≤4.3 | Ref. |  | Ref. |  |
| **Smoking status** |  |  |  |  |
| Smokers | 0.91 (0.48-1.71) | 0.766 | 0.90 (0.47-1.73) | 0.755 |
| Non-smokers | Ref. |  | Ref. |  |
| **Tuberculosis** |  |  |  |  |
| Yes | 0.68 (0.26-1.75) | 0.419 | 0.66 (0.25-1.75) | 0.408 |
| No | Ref. |  | Ref. |  |
| **Lung Function Status** |  |  |  |  |
| Restrictive | 0.62 (0.24-1.56) | 0.307 | 0.62 (0.24-1.62) | 0.333 |
| Mixed | 0.21 (0.05-0.98) | 0.046 | 0.25 (0.05-1.18) | 0.080 |
| Obstructive | 0.64 (0.26-1.62) | 0.643 | 0.61 (0.24-1.56) | 0.299 |
| Asthma | 0.32 (0.08-1.31) | 0.114 | 0.41 (0.10-1.69) | 0.216 |
| Normal | Ref. |  | Ref. |  |
| **HIV status** |  |  |  |  |
| Positive | 1.91 (0.38-9.59) | 0.433 | 1.97 (0.38-10.22) | 0.418 |
| Negative | Ref. |  | Ref. |  |
| **Diabetes Mellitus** |  |  |  |  |
| Yes | 0.88 (0.33-2.31) | 0.795 | 0.92 (0.34-2.50) | 0.874 |
| No | Ref. |  | Ref. |  |

\*The variable of population i.e. SSM and PMC was omitted since none of the participant from the PMC was found to have evidence of silicosis.

Table 5 shows the crude and adjusted odds ratios between ILF and a set of predictors. Mine workers had more than 2-times higher odds of having ILF compared to the PMC, aOR 2.06, CI=1.38-3.07, p<0.001.

**Table 5.** Factors Associated with Impaired Lung Function Among Mining Communities in Mererani (N = 660)

| Variable | cOR (95%CI) | p-value | aOR (95%CI) | p value |
| --- | --- | --- | --- | --- |
| **Population** |  |  |  |  |
| Miners | 1.90 (1.37-2.64) | 0.000 | 2.06 (1.38-3.07) | 0.000 |
| Peri-mining community | Ref. |  | Ref. |  |
| **Working duration in Mererani (years)** |  |  |  |  |
| <15 | 0.86 (0.51-1.46) | 0.584 | 0.69 (0.40-1.20) | 0.186 |
| ≥15 | Ref. |  | Ref. |  |
| **Income (USD/day)** |  |  |  |  |
| >4.3 | 0.94 (0.67-1.32) | 0.734 | 1.20 (0.83-1.75) | 0.333 |
| ≤4.3 | Ref. |  | Ref. |  |
| **Smoking status** |  |  |  |  |
| Smokers | 1.67 (1.07-2.61) | 0.024 | 1.52 (0.96-2.40) | 0.077 |
| Non-smokers | Ref. |  | Ref. |  |
| **Tuberculosis** |  |  |  |  |
| Yes | 1.41 (0.77-2.60) | 0.266 | 1.49 (0.80-2.79) | 0.209 |
| No | Ref. |  | Ref. |  |
| **Silicosis** |  |  |  |  |
| Silicosis | 0.65 (0.42-1.00) | 0.050 | 1.11 (0.68-1.79) | 0.686 |
| No silicosis | Ref. |  | Ref. |  |
| **Diabetes Mellitus** |  |  |  |  |
| Yes | 0.94 (0.46-1.90) | 0.858 | 0.734 (0.35-1.53) | 0.407 |
| No | Ref. |  | Ref. |  |

## Discussion

In light of concerns of high rates of silicosis and ILF, our study aimed to investigate the prevalence of, and risk factors for, silicosis and ILF amongst small scale tanzanite mining community i.e. both miners and PMC. In our study we found that 99/330 (30.0%) of miners had evidence of silicosis, of whom 97.0% had accelerated silicosis. The prevalence of silicosis declined with duration of working in the mines. Among miners and community members, 75 (11.4%) had ILF, of whom 30 (4.5%) had COPD, 9 (1.4%) had asthma, 29 (4.4%) had restrictive lung disease and 7 (1.1%) had mixed pattern of both obstructive and restrictive lung disease. Of the 75 participants found to have ILF, 48 (64%) were mine workers. In our analysis, we found that having a daily income of more than USD 4.3 was associated with lower odds of silicosis (aOR 0.57, 95% CI 0.37-0.89, p<0.05) while ILF was found to have a significant statistical association with occupation with miner having higher odds of ILF (aOR 2.06, CI=1.38-3.07, p<0.001).

As would be expected from pattern of occupational exposure to respirable dust between these two communities, evidence of silicosis was found only among the SSM who are constantly working in congested underground mines and not among the PMC. Due to the strong winds with frequent change of direction on daily basis (because of low vegetation) in the Mererani area, and the practice of leaving the remains of mining rocks on open grounds, we had hypothesised that the PMC may be exposed, however our recent exposure data [10] and this study suggest this not to be the case. Though the current study sampled only the drillers but as reported by the miners, there is frequent, almost on daily basis, such that any miner can be allocated to undertake any tasks between drilling, blasting, and shovelling. But generally, at any given time around 50% of the miners will be involved with drilling, around 40% involved with shovelling and around 10% work on blasting.

Reports have shown that short periods of intense exposure to RCS to be associated with significantly higher risk of diseases, including silicosis [21]. This could be the case with the findings from the current study in which those who worked for less than 15 years (consistent with accelerated silicosis) had more than twice higher odds of evidence of silicosis relative to miners who have worked for at least 15years (though the finding was not statistically significant). However, the current study finding of most (97%) of those with evidence of silicosis to have worked for a duration of less than 15years, could otherwise be explained by the “healthy worker effect”. That as the severity of disease increases with time, exposed workers become too ill, possibly unable to go down the pits anymore, and leave mining work to seek health services (usually at their home residencies) or to change to other kind of occupation out of the mines. A similar pattern of silicosis prevalence has been observed among small-scale gold miners in China [8]. Our study showed a silicosis prevalence of 30%, which is towards the higher end though in keeping with other studies of SSM [9]. Kilimanjaro Christian Medical Centre (KCMC) in Moshi, Tanzania has been managing patients from Mererani mines for years. A retrospective hospital-based data analysis among patients admitted due respiratory conditions at KCMC from August 2010 to August 2020, revealed the prevalence of silicosis of 14.3% [22], about half of what has been reported from the current study. Studies among other populations have reported a lower prevalence, including that among stone crushers in Haryana, India of around 6.4% [23] and gold miners in South Africa 19.9% [24]. Our selection criteria meant that current study participants were all engaging in drilling of rocks (though in real practice, there is task shifting in which most of the workers will do different work at varying times, including blasting and shovelling of rocks) and given the complete absence of dust reduction mechanisms, including not using water spray during drilling and poorly ventilated mining pits, it could explain this high level of silicosis observed. Our finding of high prevalence is complemented by the findings of high levels of exposure to respirable crystalline silica (RCS) among the same current study participants [10]. Our study may also under-estimate the true prevalence of disease. A study conducted in China among coal workers reported an almost 27% more patients with Coal Workers Pneumoconiosis (CWP) when examined by High Resolution Computed Tomography (HRCT) scan as compared to examination done by Film-Screen Radiography [25]. This difference signifies the variation that may arise because of different diagnostic techniques, hence the possibility of an even higher burden of silicosis finding in the current study if HRCT was used.

The current study finding of evidence of silicosis being associated with income level could be explained by idea that miners who, for different reasons, have higher income (being it from the mining work or other sources) usually spend less time underground compared to the rest of the miners, hence less exposed to RCS and therefore less chances of developing silicosis. The SSM don’t have regular salaries, they have entered into agreement with mine managers to get a certain percentage, usually around 5% of the sales to share among all the workers once a gemstone is found. In contrast to a recent study in Rutsiro, Rwanda [26] average earnings in Mererani were relatively low with the majority of miners on <4.3 USD/day. In the Rwandan study, reported SSM to have not only regular monthly salaries but the salary being relatively good to the extent of attracting workers from other sectors like construction to the mining sector. Among these SSM interviewed about ill health related to the mining activities, 85% reported dust to be a key challenge, of whom 65% reported silicosis as a major health challenge [26]. While improving workers’ income should be prioritized, this should go hand in hand with improvement of the working environment to ensure the individuals who are rushing in the mines for better paying jobs are in a safe working condition.

The current study report 11.4% of the study participants to have ILF comprising of COPD, Asthma, RLD or mixed OLD/RLD). The prevalence of ILF appears to be slightly higher among miners than the PMC, although the study was likely underpowered to investigate this fully. The higher prevalence of ILF among miners could be associated with the occupational exposure to respirable dust. Our group has already demonstrated very high exposures to respirable crystalline silica among miners, and low exposures among PMC [10]. It may also be related to a higher prevalence of smoking (18% vs 11%) among the miners compared to the PMC. The prevalence of ILF reported in our study is broadly similar to those reported in other studies. Retrospective cross-sectional analysis of hospital data at KCMC in Moshi, Tanzania showed COPD and asthma to rank the fourth and sixth respectively as common diseases among patients admitted with respiratory conditions (that include SSM from Mererani), though mining accounted for just a small percentage among those with CODP and none among those with asthma [22]. A pilot study to assess respiratory health among community residing near to gold mines waste in Johannesburg South Africa which defined levels of exposures based on distance from the mine waste as high (home <500m), moderate (500m–1.5km) and low (>1.5km), showed the prevalence of COPD to be more common in the high exposure group (18.6%) relative to the medium exposure (10.2%) and low exposure (10.6%) groups [27]. Compared to the current study, the PMC (located about 5km from the tanzanite mines) will belong to the low exposure group residing > 1.5km from the mines’ waste. A study report among non-smokers coal workers (both underground and surface) in USA showed the prevalence of obstructive lung disease of 7.7% which went up to 16.4% among those with CWP [28] while another report among miners in Western Australia reported a prevalence of 6.3% [29].

A univariate analysis of association for a study conducted in China reported COPD to be associated with exposure to silica dust, pneumoconiosis stage III and heavy cigarette smoking while on multi-variate analysis of association, COPD was found to be associated with exposure to silica dust and pneumoconiosis stage III [17]. In addition, [30] reported FEV_1_ and FVC to be lower among smokers compared to non-smokers. Though the analysis of association in the current study did not segregate the ILF into the sub-types, but smoking was found to be associated with ILF with about 1.7 higher odds of ILF among the smokers. Unlike the current study findings that did not show an association between ILF and duration of work, a study report among iron ore workers in Iran reported duration of work to significantly lower the FVC, FEV_1_ and FEV_1_/FCV [31]. Opposite findings have been reported from a comparison cross-sectional study among stone crushers in Democratic Republic of Congo (DRC), in which the stone crushers (exposed group) had higher FVC and FEV_1_ compared to the tax drivers, the unexposed group [32] and also a study among underground gold miners in Tanzania reported that smoking characteristics and history of dust exposure to be not associated with ILF [18].

Mine workers with history of suffering from TB in South Africa were reported to have reduced lung function with FEV_1_ of about 180mls-lower and FVC of about 120mls-lower relative to those who never suffered TB [33]. As reported in our previous study report [15], the prevalence of TB among the mining communities in Mererani was around 7.0% which is about 18-times higher compared to the national prevalence of around 0.4% (WHO TB burden estimates 2021; https://www.who.int/teams/global-tuberculosis-programme/data). Unexpectedly, the TB burden was higher (7.9%) in the PMC compared to 6.1% for SSM. Given the high burden of TB in both SSM and PMC, it is likely that the high prevalence (53.2%) of previous lung disease found in the current study (Table 1), was contributed by previous TB. As has been shown by [34] in their study conducted among adult who have completed TB treatment in Kilimanjaro Tanzania to determine the burden and severity of post tuberculosis lung disease (PTLD), the prevalence of PTLD was found to be 91%.

Limitations in this study included: not utilizing CT scan, which is more sensitive than chest digital X-ray in assessing the pulmonary disease, hence could have missed some of the radiological features associated with silicosis. In addition, for the PMC the specific type of an individual’s work (aside from being non-miner) was not determined, which could have allowed comparison of investigated diseases in different occupational situations. Only one radiologist read the digital chest X-rays, though has more than 11 years’ experience in the field of radiology, including occupational related diseases like silicosis. In addition, using a cross-sectional design could have underestimated the burden of the evaluated diseases.

Silicosis is a major health concern among the SSM and ILF is highly prevalent among both SSM and the PMC in Mererani, in northern Tanzania. As silicosis is an incurable but potentially preventable condition, immediate dust control measures including deployment of wet drilling, wearing of PPE and regular monitoring of dust exposure need to be enforced by OSHA (Tanzania). As per the Occupational Safety and Health Act of 2003 [35], the Chief Inspector should take appropriate measures to mine owners not abiding the safety and health measures. To facilitate this, Tanzania should join the ILO/WHO global efforts to end silicosis by 2030. Health facilities, especially those serving the mining communities should establish and strengthen expertise in the management of chronic lung diseases, especially COPD. Longitudinal studies focusing on wider aspects of occupational exposure to hazardous materials and their disease implications should be undertaken in this community.

## Data Availability

Data have been provided as part of the submitted article

## Acknowledgements

I would like to acknowledge all professors, tutors, and lecturers from the KCMUCo Institute of Public Health for their precious time on imparting new knowledge on me and making close follow up on my academic progress. We would like to thank the LHL International (Oslo, Norway) for their support on all the health services provided to the patients, including transportation to/from the clinic where the investigations were conducted. Our sincere thanks for staff of OHSC at Kibong’oto Hospital for their support and much of the clinical work.

## Supporting information captions

1. S1 Fig. 1. Duration of work in Mererani mines among mine workers found to have evidence of silicosis.
2. S2 Fig. 2. Scatter plot showing the spread of various forms of lung function status as functions of FVC and FEV1.
3. S3 Text. Interview schedule.
4. S4 Text. Consent form.
5. S5 Data. All collected data

